# Effects of Extra-curricular Physical Activity Programs on High-school Girls: A Systematic Review

**DOI:** 10.1101/2020.04.25.20079780

**Authors:** Laura-Maude Houle, Jo-Anne Gilbert, Karine Paiement, Alexandra Ayotte, Marie-Eve Mathieu

## Abstract

**Introduction:** Most adolescents do not meet physical activity (PA) recommendations, especially girls. Physical inactivity has major physical and psychosocial deleterious effects on adolescents. Little is known about the effect of girl-only, extra-curricular PA programs designed for adolescents on physical and psychosocial outcomes. Hence, this systematic review assessed quantitative and qualitative studies evaluating the effects of such interventions. It also aimed at identifying recommendations to improve their implementation and efficacy.

**Methods:** Literature published until June 4, 2018, was searched in three electronic databases. Two reviewers independently assessed the methodological quality of studies presenting results on physical and psychosocial outcomes, not those presenting solely implementation recommendations.

**Results:** Seventeen quantitative and qualitative studies describing 10 different PA programs were included. Six of these studies provided recommendations for implementation only. The overall quality of the studies assessing the effects on physical and psychosocial outcomes was moderate, with an average score of 58%. The PA programs assessed did not lead to clear improvements in PA levels or other physical outcomes. Concerning psychosocial results, there is some evidence that the programs could improve dimensions of the self-esteem construct.

**Conclusion:** Future studies should assess the effect of girl-only, extra-curricular PA programs on the health-related habits, such as reduction of sedentary time among adolescents and sleep duration. More studies evaluating the psychosocial effects as a primary outcome are recommended to obtain a clearer understanding of the benefits. This review gathers recommendations to improve the efficacy of future extra-curricular PA programs designed to increase PA levels in girls.

## Introduction

Important developmental changes occur during adolescence and an active lifestyle is beneficial for physical and mental health and well-being throughout this life stage. Evidence supports the impact of physical activity (PA) on the prevention of obesity, cardiovascular disease, metabolic syndrome and osteoporosis in children and adolescents (Janssen & LeBlanc, 2010). Moreover, PA can improve mental-health (Korczak, Madigan, & Colasanto, 2017) as well as cognitive and academic performance (Li, O’Connor, O’Dwyer, & Orr, 2017). However, three-quarters of adolescents worldwide do not meet the daily PA recommendation (World Health Organization, 2018a). Insufficient PA participation is more prevalent among adolescent girls (84%) than adolescent boys (78%) (World Health Organization, 2018b). While both sexes experience a decline in PA levels over time during adolescence, the decline is more pronounced in girls (Dumith, Gigante, Domingues, & Kohl, 2011).

School settings offer great opportunities to promote the adoption of an active lifestyle. A recent meta-analysis of PA interventions showed that school-based interventions had a greater effect than community-based programs on the PA levels of adolescent girls (Pearson, Braithwaite, & Biddle, 2015). However, there is room for improvement to make sure adolescent girls benefit from school-based PA programs. First, most programs do not address the specific barriers reported by adolescent girls, such as the lack of variety of activities, a competitive environment and the absence of friends (Martins, Marques, Sarmento, & Carreiro da Costa, 2015). The presence of boys was also reported as a barrier. However, the majority of school PA is mixed. In this regard, there is some evidence that girl-only PA interventions are more successful for adolescent girls than mixed-sex interventions (Pearson et al., 2015). Second, many school-based PA interventions focus solely on enhancing physical education classes. Knowing that PA levels in secondary school physical education are often far below the national recommendation, it seems interesting to increase PA levels during the classes (Hills, Dengel, & Lubans, 2015). However, there is generally only a few hours of class per week, limiting their potential to influence total PA participation (Owen, Curry, Kerner, Newson, & Fairclough, 2017). In this context, extra-curricular programs run in school settings provide additional opportunities to be active for adolescent girls, especially when the interventions address their particular barriers to PA.

There is only few evidences on the effects of extra-curricular PA programs (ECPAP) on adolescents and even less on adolescent girls. For instance, a review performed in 2009 suggested that after-school PA programs could increase PA levels and some health-related outcomes in youth, but the authors did not make distinctions for sex and life stage (Beets, Beighle, Erwin, & Huberty, 2009). Subsequent reviews assessed girls PA participation in response to school-based programs, but did not evaluate separately child and adolescent girls and did not isolate the effects of programs developed especially for girls (Camacho-Minano, LaVoi, & Barr-Anderson, 2011; Voskuil, Frambes, & Robbins, 2017). The most recent review published on the topic assessed all school-based PA interventions for adolescent girls but did not single out the effects of ECPAP. To our knowledge, the current systematic review is the first to assess the efficacy of ECPAP specifically targeting adolescent girls. Since most reviews reported only the effects on PA levels, the objective of the present review is to evaluate the effects of ECPAP on various physical and psychological outcomes. It also aims to report the recommendations for successful implementation and effectiveness of such programs.

## Methods

This systematic review follows the procedures of the preferred reporting items for systematic reviews and meta-analyses PRISMA guidelines (Moher, Liberati, Tetzlaff, & Altman, 2009) (Appendix A).

### Eligibility criteria

To be eligible, studies needed to involve adolescent girls aged 11 to 17 years attending high school. We included studies with a sample mean age in this range and those with at least some participants attending high school. Studies assessed a single-sex PA intervention developed specifically for adolescent girls. The PA component of the intervention was implemented in a school setting, during the school calendar year, but outside the teaching hours, such as before/after school or during the lunch break. Schools needed to be involved in either the implementation or facilitation of the PA intervention. Because the first objective of this review is to understand the effects of ECPAP, we included studies reporting physical or psychological outcomes, such as PA levels, fitness markers, and self-esteem levels. Because it also aims to gather recommendations for successful implementation and effectiveness of ECPAP, studies assessing programs’ characteristics were included. Hence, this systematic review contains a wide range of intervention and qualitative studies that included or not a control group. The presence and length of follow-up assessments were not part of the inclusion/exclusion criteria. Only papers published in English in scientific journals were retained for this review. Studies were excluded if 1) they targeted populations with diagnosed illness, diseases or disability; 2) the intervention was run exclusively during physical education classes; 3) they had no relevant results to extract, such as observational studies, comments and opinions articles as well as those solely describing research design (methods, protocols, theories).

### Literature search strategy

We searched literature published until June 4, 2018, with no limitation on the start date. The search was conducted using three English-language electronic databases (ERIC, Sport Discuss, Web of Science). The following keywords were used: (athletics, dance exercise, physical activities, physical activity, sport and/or workout) AND (after school, after school, before school, curricular, co-curricular, enrichment, extended school day, extracurricular, extra curriculum, intercollegiate, interscholastic, intramural, lunch break, lunch recess, lunchtime, lunchtime, non-school hours, out-of-school, recreational and/or school after hours), AND (adolescent, teen, youth, college, high school and/or secondary school). The search strategy was specific to each database. To capture all relevant studies, pilot searches were carried out to test the search filters and ensure the quality of the final search strategy.

### Screening

Two reviewers (LMH, KP) independently determined study’s eligibility. Both reviewers identified potentially relevant articles through title and abstract screening. The inter-rater reliability of the initial screening of the manuscripts was determined by two reviewers with an agreement of at least 70%. Any disagreements between the two reviewers was discussed and resolved by consulting a third reviewer (MEM) raising the agreement cut-off to 85%. After discussion, the three reviewers obtained 100% agreement. Finally, the first two reviewers looked over studies full text to determine studies to be included in the systematic review.

### Data extraction and analysis

The following information was recorded in a table:

▪ First author, year and country
▪ Sample characteristics (e.g. sample size and ethnic/racial group of the participants, age of participants)
▪ Objective
▪ Intervention description
▪ Study design
▪ Physical and psychosocial outcomes
▪ Main findings
▪ Recommendations for the implementation and success of the intervention

When studies presented multiple measures of a parameter, only the most relevant measure was extracted. Inconsistencies between reviewers (LMH, KP) were resolved by an additional one (MEM) who also revised the data set. When additional information was needed, one of the reviewers contacted the corresponding authors of the original articles.

Findings from the studies were synthesized using categories: 1) physiological outcomes including PA levels, 2) psychosocial outcomes and 3) program characteristics or recommendations for improving its implementation and success rate. Since this systematic review aims to assess the various effects of ECPAP, one could expect a wide variety like the results reported. To enable the interpretation, a system indicating whether each study led to at least some positive (+), some negative (-), or no effects (0) was used. Previous PA reviews have used similar systems (Sallis, Prochaska, & Taylor, 2000). To assign the positive sign (+) to an outcome, the program had to lead to significant quantitative differences or some qualitative positive effects. The negative sign (-) was assigned using the same method. The neutral sign (0) means that no statistical differences or qualitative effects were reported.

### Methodological quality assessment

Two reviewers (KP, JAG) assessed the methodical quality of studies describing the effects of the ECPAP on physical and psychosocial outcomes. They used a checklist of 27 questions designed to assess both randomised and non-randomised trials to consider the nature of the studies included (Downs & Black, 1998) (Appendix B). It assesses the risk of bias coming from the reporting of information, external and internal validation, and power. Some modifications were applied to improve the scoring system, as used previously (Knols, Fischer, Kohlbrenner, Manettas, & de Bruin, 2018). The scoring represents the percentage of criteria properly fulfilled. Greater percentage means higher methodological quality. The scoring was used in combination with the signs indicating positive, negative or neutral findings to refine the conclusions drawn for each physical and psychosocial outcomes. Studies that presented solely implementation recommendations were not assessed using the checklist because they did not fit the criteria that would have made them eligible for such examination. For example, power calculations were not available for qualitative studies.

## Results

Figure 1 describes the screening process performed. The publication year of the included studies started at 2000, as presented in Table 1. Many articles came from the implementation of the same ECPAP, sometimes with different populations and settings. Hence, this review included 17 studies presenting the results coming from 10 different ECPAP. As shown in Figure 1, the risk of bias was assessed for the 11 studies that reported the program’ effects on physical and/or psychological outcomes. Five studies provided only information on the best practice for program implementation (Edwards et al., 2016; R. Jago et al., 2012; Rajan & Basch, 2012; Robbins, Ling, Toruner, Bourne, & Pfeiffer, 2016; Sebire et al., 2016).

**Table 1.**
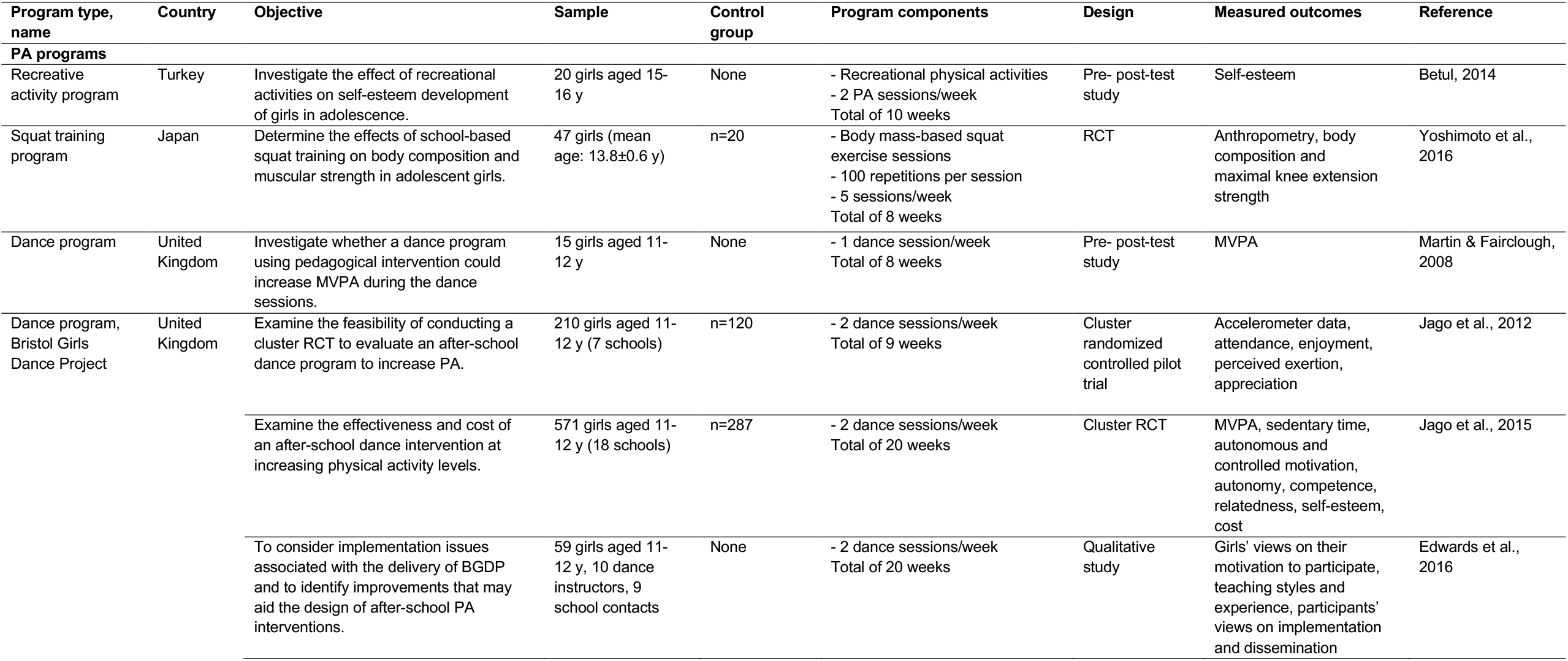
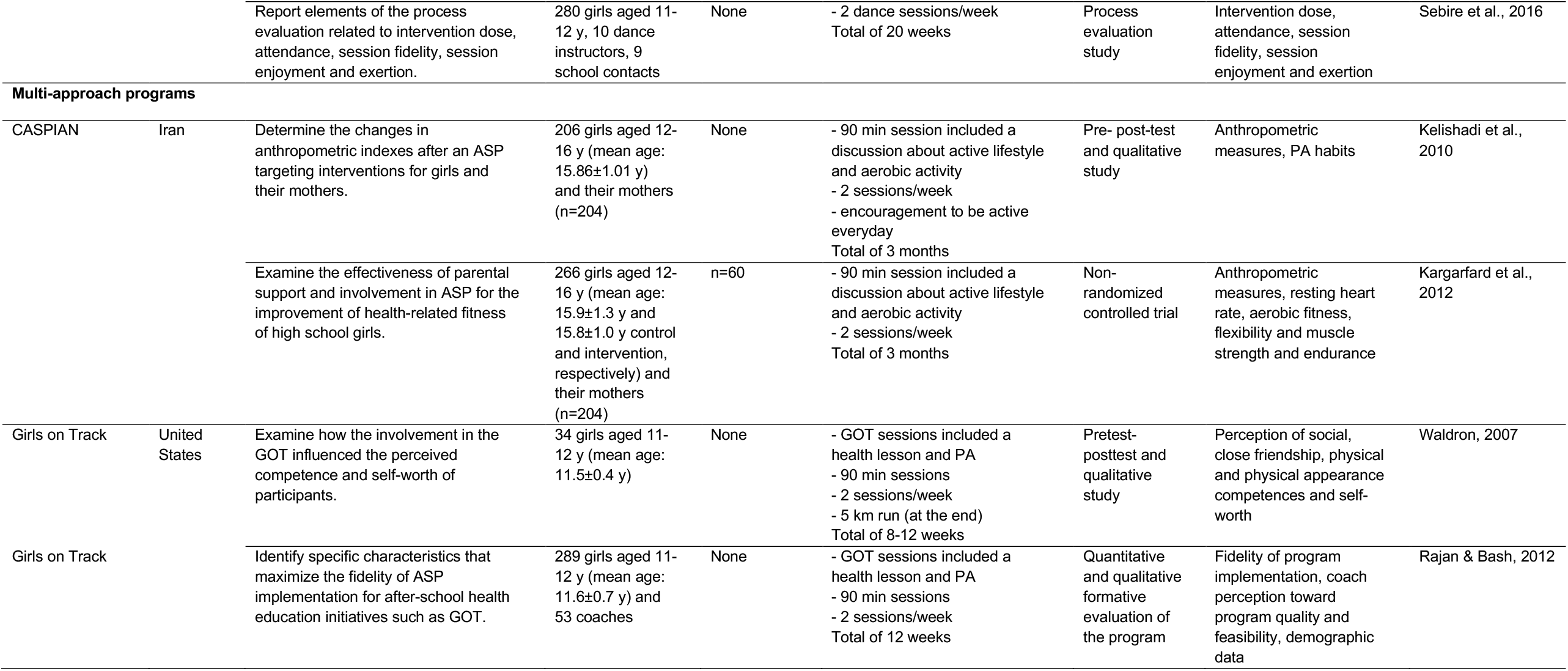
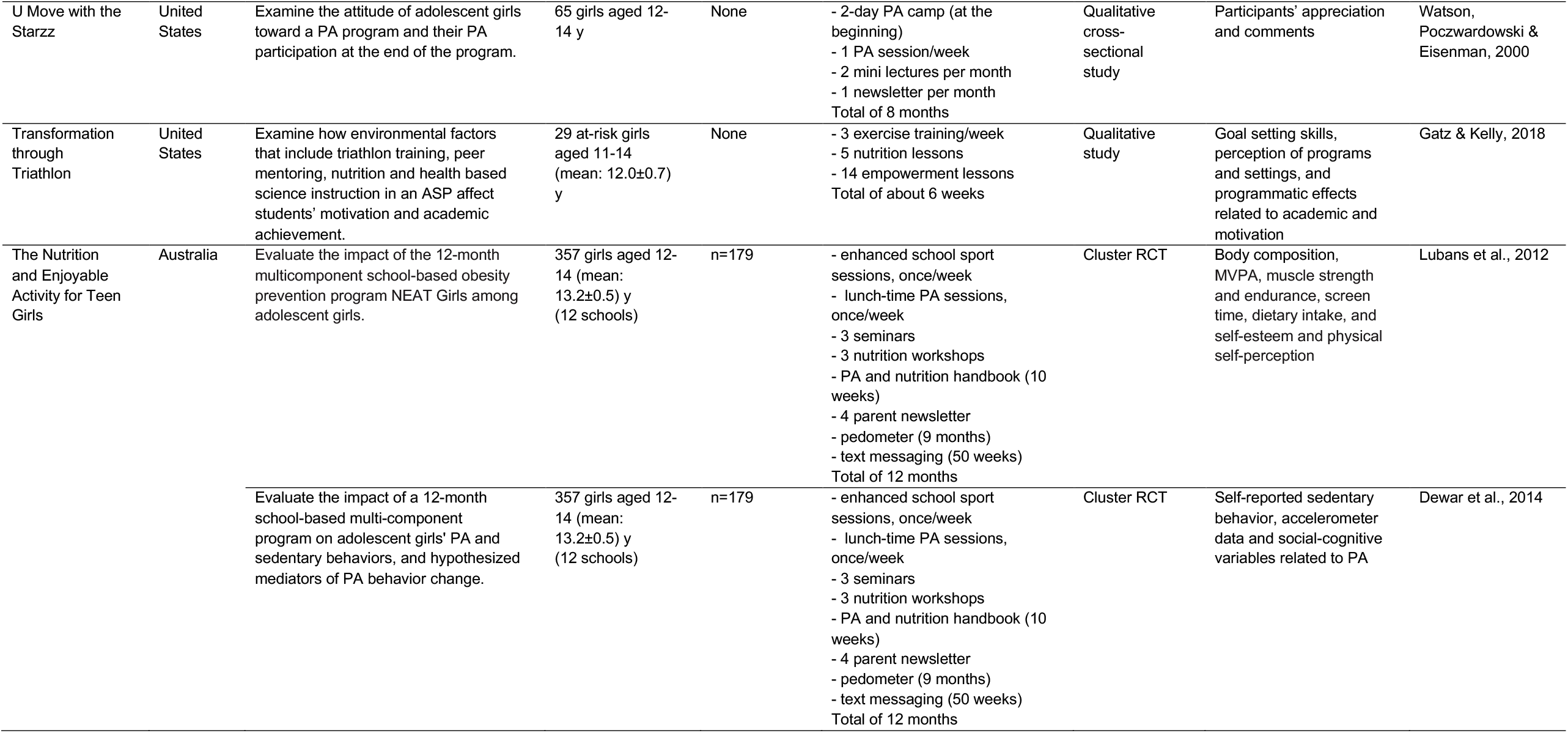
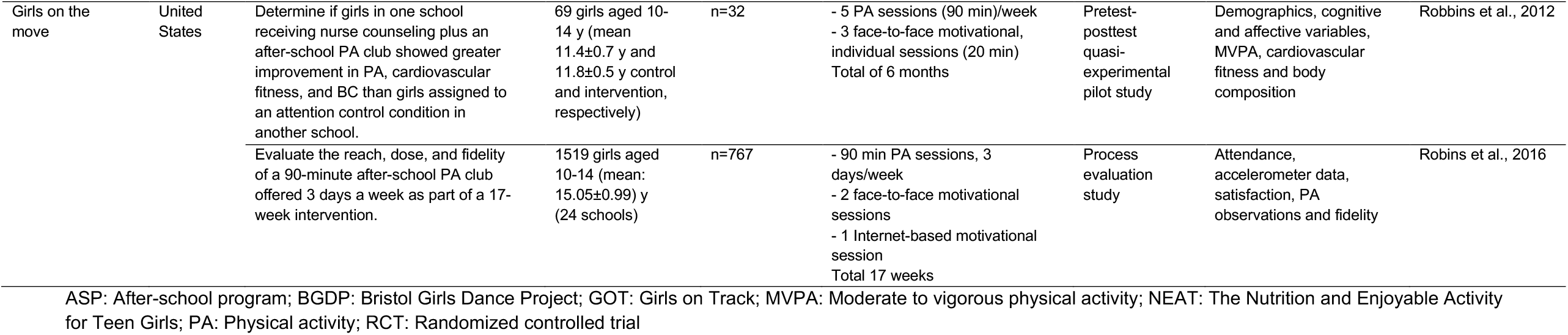
Programs included in the systematic review

**Figure 1:**
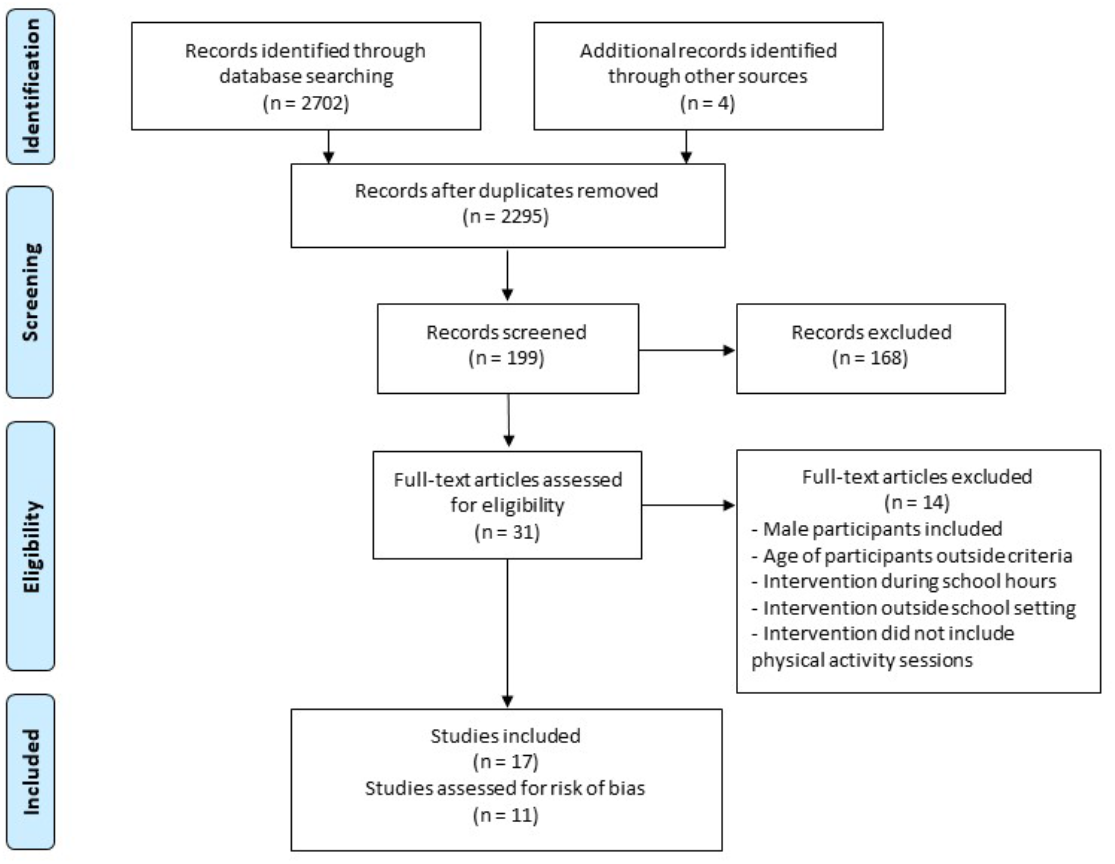
Flowchart of the literature search strategy

### Program characteristics

Table 1 shows that studies were conducted in many countries, but most were from the United States. Programs lasted between 6 weeks and 12 months. The frequency of PA sessions varied between 1 and 5 sessions per week, most ECPAP (6 out of 10) included 2 or 3 sessions weekly. The number of participants varied between 15 and 1519 adolescent girls. Most studies included adolescents in the younger age range. Four ECPAP used PA as the only component of the intervention while the six others used additional components. In the interventions where PA was the only component, the authors mentioned the nature of the activity, such as dance or squat training. However, in multi-approach programs, the PA intervention was rarely described. Overall, the authors provided very little details on the intensity of the PA intervention.

Among the four ECPAP based solely on PA intervention, three used only one discipline. Two of these ECPAP were dance programs (Edwards et al., 2016; Jago et al., 2015; Jago et al., 2012; Sebire et al., 2016) and one used squat training (Yoshimoto et al., 2016). The other ECPAP included multiple recreational sports and activities (Betul, 2014).

All six multi-approach ECPAP included some discussions either one-on-one or in groups (Table 1). Discussions covered one or many of the following topics: health, PA, nutrition, empowerment and skill development. The group discussions were in the form of led discussions, seminars, lessons or workshops. Some multi-approach ECPAP included a motivational component. One study included an internet-based motivational session (Robbins et al., 2016). The CASPIAN program was the only program that added parental participation in PA as a motivational component for the adolescent girls (Kargarfard et al., 2012; Kelishadi et al., 2010). One ECPAP sent monthly newsletters (Watson, Poczwardowski, & Eisenman, 2000) and one combined monthly newsletters and regular encouraging text messages (Dewar et al., 2014; Lubans et al., 2012).

As shown in Table 1, the majority of the studies reported quantitative data that were collected mostly using self-reported questionnaires, anthropometric measures, and accelerometers. The studies that collected qualitative data used interviews and focus groups (Edwards et al., 2016; Gatz & Kelly, 2018; Rajan & Basch, 2012; Sebire et al., 2016; Waldron, 2007; Watson et al., 2000).

### Physical outcomes

As shown in Table 2, the physical outcomes extracted were grouped into four categories that are PA levels, sedentary activities, fitness markers, and body composition. The PA levels category presents the levels of moderate-to-vigorous physical activity (MVPA) as determined using accelerometry data. In one study only, MVPA was measured during the PA sessions (Martin & Fairclough, 2008) as opposed to the estimation of mean MVPA spent daily (Jago et al., 2015; Lubans et al., 2012; Robbins, Pfeiffer, Maier, Lo, & Wesolek Ladrig, 2012). Martin and Fairclough (2008) observed a significant increase in MVPA of 19.3 ± 1.7 min (p<0.05) during their enhanced dance classes when compared to pre-intervention classes. Using a more robust design, Jago et al. (2015) noted that the intervention increased significantly daily minutes in MVPA on intervention days only. No intervention increased in mean daily MVPA minutes when averaged over several days (Table 2). Between study heterogeneity makes impossible the comparison of the effect sizes on mean daily MVPA. There were differences in the MVPA definitions used and statistical adjustments.

**Table 2.**
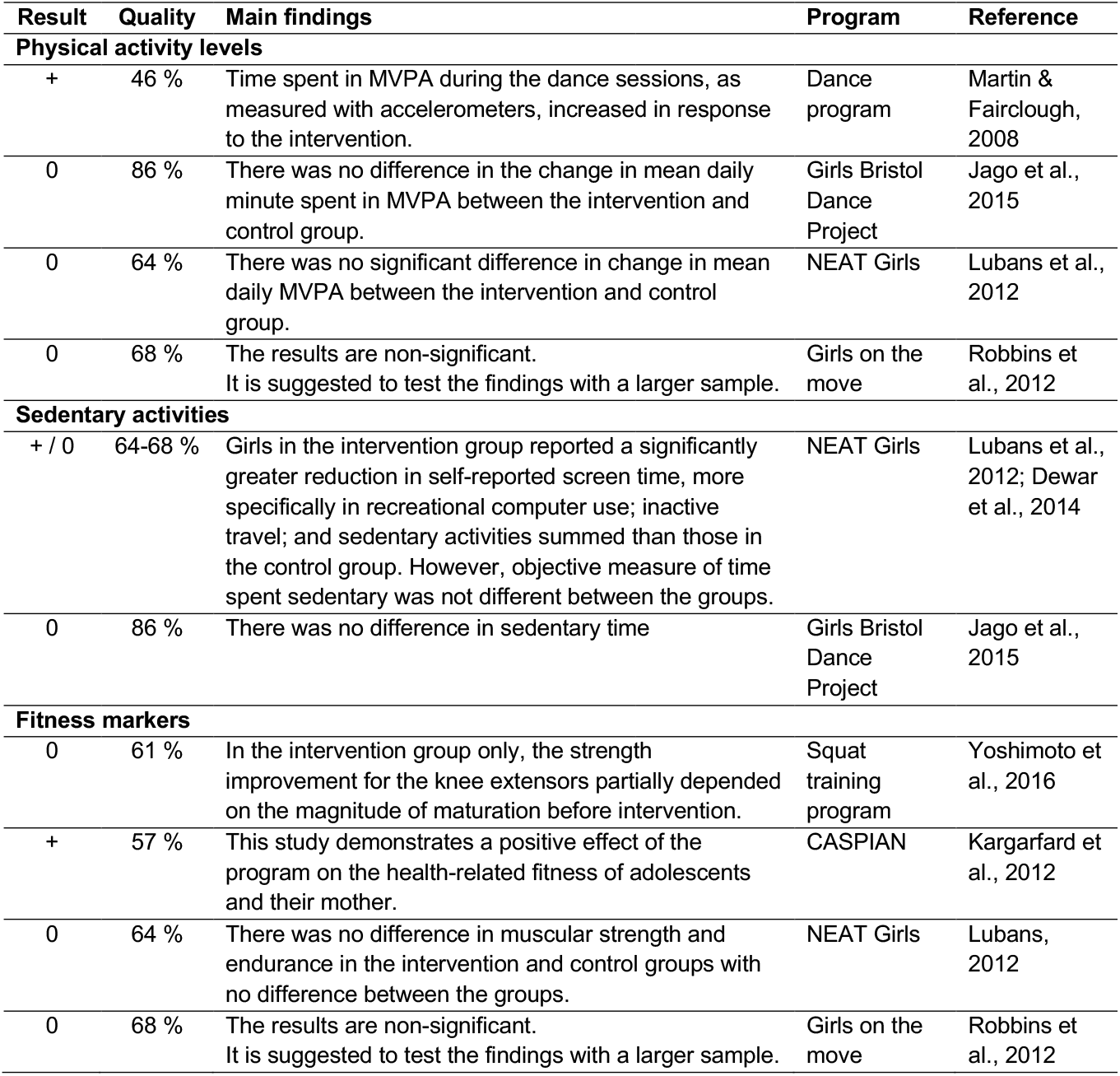
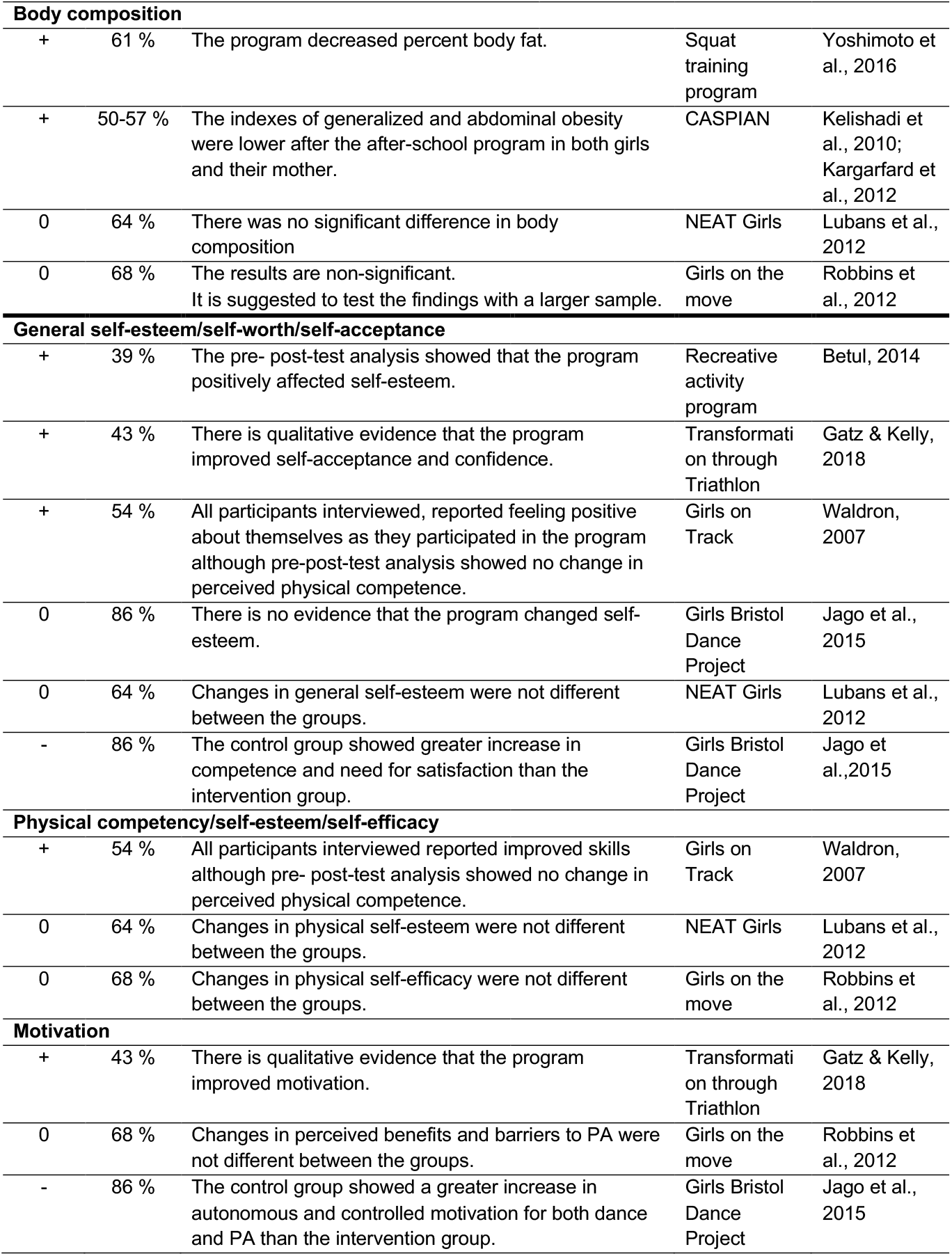
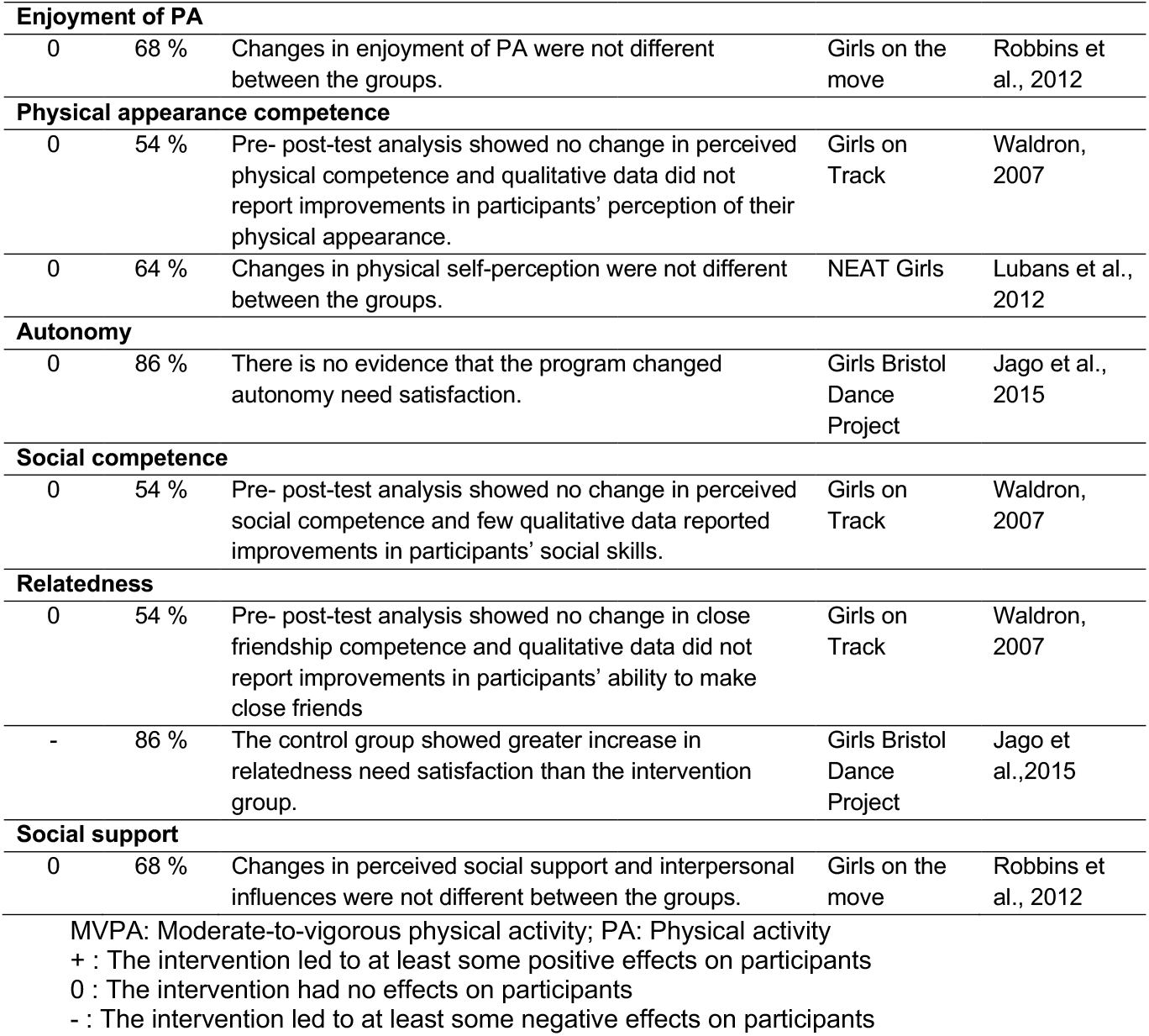
Findings on the physical and psychosocial effects of extra-curricular physical activity programs

| Result | Quality | Main findings | Program | Reference |
| --- | --- | --- | --- | --- |
| **Physical activity levels** |  |  |  |  |
| + | 46 % | Time spent in MVPA during the dance sessions, as measured with accelerometers, increased in response to the intervention. | Dance program | Martin & Fairclough, 2008 |
| 0 | 86 % | There was no difference in the change in mean daily minute spent in MVPA between the intervention and control group. | Girls Bristol Dance Project | Jago et al., 2015 |
| 0 | 64 % | There was no significant difference in change in mean daily MVPA between the intervention and control group. | NEAT Girls | Lubans et al., 2012 |
| 0 | 68 % | The results are non-significant. It is suggested to test the findings with a larger sample. | Girls on the move | Robbins et al., 2012 |
| **Sedentary activities** |  |  |  |  |
| + / 0 | 64-68 % | Girls in the intervention group reported a significantly greater reduction in self-reported screen time, more specifically in recreational computer use; inactive travel; and sedentary activities summed than those in the control group. However, objective measure of time spent sedentary was not different between the groups. | NEAT Girls | Lubans et al., 2012; Dewar et al., 2014 |
| 0 | 86 % | There was no difference in sedentary time | Girls Bristol Dance Project | Jago et al., 2015 |
| **Fitness markers** |  |  |  |  |
| 0 | 61 % | In the intervention group only, the strength improvement for the knee extensors partially depended on the magnitude of maturation before intervention. | Squat training program | Yoshimoto et al., 2016 |
| + | 57 % | This study demonstrates a positive effect of the program on the health-related fitness of adolescents and their mother. | CASPIAN | Kargarfard et al., 2012 |
| 0 | 64 % | There was no difference in muscular strength and endurance in the intervention and control groups with no difference between the groups. | NEAT Girls | Lubans, 2012 |
| 0 | 68 % | The results are non-significant. It is suggested to test the findings with a larger sample. | Girls on the move | Robbins et al., 2012 |
| **Body composition** |  |  |  |  |
| + | 61 % | The program decreased percent body fat. | Squat training program | Yoshimoto et al., 2016 |
| + | 50-57 % | The indexes of generalized and abdominal obesity were lower after the after-school program in both girls and their mother. | CASPIAN | Kelishadi et al., 2010; Kargarfard et al., 2012 |
| 0 | 64 % | There was no significant difference in body composition | NEAT Girls | Lubans et al., 2012 |
| 0 | 68 % | The results are non-significant.<br>It is suggested to test the findings with a larger sample. | Girls on the move | Robbins et al., 2012 |
| **General self-esteem/self-worth/self-acceptance** |  |  |  |  |
| + | 39 % | The pre- post-test analysis showed that the program positively affected self-esteem. | Recreative activity program | Betul, 2014 |
| + | 43 % | There is qualitative evidence that the program improved self-acceptance and confidence. | Transformati on through Triathlon | Gatz & Kelly, 2018 |
| + | 54 % | All participants interviewed, reported feeling positive about themselves as they participated in the program although pre-post-test analysis showed no change in perceived physical competence. | Girls on Track | Waldron, 2007 |
| 0 | 86 % | There is no evidence that the program changed self-esteem. | Girls Bristol Dance Project | Jago et al., 2015 |
| 0 | 64 % | Changes in general self-esteem were not different between the groups. | NEAT Girls | Lubans et al., 2012 |
| - | 86 % | The control group showed greater increase in competence and need for satisfaction than the intervention group. | Girls Bristol Dance Project | Jago et al., 2015 |
| **Physical competency/self-esteem/self-efficacy** |  |  |  |  |
| + | 54 % | All participants interviewed reported improved skills although pre- post-test analysis showed no change in perceived physical competence. | Girls on Track | Waldron, 2007 |
| 0 | 64 % | Changes in physical self-esteem were not different between the groups. | NEAT Girls | Lubans et al., 2012 |
| 0 | 68 % | Changes in physical self-efficacy were not different between the groups. | Girls on the move | Robbins et al., 2012 |
| **Motivation** |  |  |  |  |
| + | 43 % | There is qualitative evidence that the program improved motivation. | Transformati on through Triathlon | Gatz & Kelly, 2018 |
| 0 | 68 % | Changes in perceived benefits and barriers to PA were not different between the groups. | Girls on the move | Robbins et al., 2012 |
| - | 86 % | The control group showed a greater increase in autonomous and controlled motivation for both dance and PA than the intervention group. | Girls Bristol Dance Project | Jago et al., 2015 |
| **Enjoyment of PA** |  |  |  |  |
| 0 | 68 % | Changes in enjoyment of PA were not different between the groups. | Girls on the move | Robbins et al., 2012 |
| **Physical appearance competence** |  |  |  |  |
| 0 | 54 % | Pre- post-test analysis showed no change in perceived physical competence and qualitative data did not report improvements in participants' perception of their physical appearance. | Girls on Track | Waldron, 2007 |
| 0 | 64 % | Changes in physical self-perception were not different between the groups. | NEAT Girls | Lubans et al., 2012 |
| **Autonomy** |  |  |  |  |
| 0 | 86 % | There is no evidence that the program changed autonomy need satisfaction. | Girls Bristol Dance Project | Jago et al., 2015 |
| **Social competence** |  |  |  |  |
| 0 | 54 % | Pre- post-test analysis showed no change in perceived social competence and few qualitative data reported improvements in participants' social skills. | Girls on Track | Waldron, 2007 |
| **Relatedness** |  |  |  |  |
| 0 | 54 % | Pre- post-test analysis showed no change in close friendship competence and qualitative data did not report improvements in participants' ability to make close friends | Girls on Track | Waldron, 2007 |
| - | 86 % | The control group showed greater increase in relatedness need satisfaction than the intervention group. | Girls Bristol Dance Project | Jago et al., 2015 |
| **Social support** |  |  |  |  |
| 0 | 68 % | Changes in perceived social support and interpersonal influences were not different between the groups. | Girls on the move | Robbins et al., 2012 |
| MVPA: Moderate-to-vigorous physical activity; PA: Physical activity<br>+ : The intervention led to at least some positive effects on participants<br>0 : The intervention had no effects on participants<br>- : The intervention led to at least some negative effects on participants |  |  |  |  |

The sedentary activities were reported for two ECPAP. The NEAT Girls program reduced significantly self-reported screen time, especially recreational computer use, and the sum of sedentary activities, with an adjusted median difference (interquartile) of −30.7 (−62.4 to −1.06) min/d, −26.0 (−46.9 to −5.1) min/d, and −56.4 (−110.1 to −2.7) min/d, respectively (Dewar et al., 2014; Lubans et al., 2012). However, the objective measure of time spent sedentary using accelerometry did not show differences between the intervention and control group in both ECPAP (Dewar et al., 2014; Jago et al., 2015).

The fitness markers category included various measures, that is muscle strength (Kargarfard et al., 2012; Lubans et al., 2012; Yoshimoto et al., 2016), aerobic fitness (Kargarfard et al., 2012; Robbins et al., 2012) and flexibility (Kargarfard et al., 2012). The study from Kargarfrard et al. (2012) was the only one to report any improvement in fitness markers. After the 12-week intervention, participants significantly increased their maximal oxygen consumption, flexibility, and abdominal muscular strength when compared to baseline.

Body composition outcomes came from measured data exclusively. Outcomes were percent body fat (Lubans et al., 2012; Robbins et al., 2012; Yoshimoto et al., 2016), body mass index (BMI) (Kargarfard et al., 2012; Kelishadi et al., 2010; Lubans et al., 2012; Robbins et al., 2012; Yoshimoto et al., 2016), BMI z-score (Lubans et al., 2012; Robbins et al., 2012), BMI percentile for age (Robbins et al., 2012), waist circumference (Kelishadi et al., 2010; Robbins et al., 2012), and waist-to-hip ratio (Kelishadi et al., 2010). The longer interventions, that lasted 6 months (Robbins et al., 2012) and 12 months (Lubans et al., 2012), did not lead to changes in body composition. The shorter ones, lasting 8 weeks (Yoshimoto et al., 2016) and 12 weeks (Kargarfard et al., 2012; Kelishadi et al., 2010), resulted in reduced markers of adiposity.

### Psychosocial outcomes

The evaluation of psychosocial outcomes was the primary objective of three studies (Betul, 2014; Gatz & Kelly, 2018; Waldron, 2007), while it was among the secondary objectives for the others (Table 2). Most studies used quantitative questionnaires as measuring tools and two studies used qualitative measures. Waldron (2007) selected eight participants among those who completed the ECPAP to take part in semi-structured interviews. These interviews complemented the quantitative analyses performed using the whole sample (Waldron, 2007). Gatz and Kelly (2018) used solely focus groups to evaluate the effect of their ECPAP. Many psychosocial constructs have been measured using various evaluation tools. For the purpose of this review, constructs have been regrouped in broad concepts to make their interpretation simpler (Table 2).

At least some positive effects have been reported for general and physical self-esteem as well as motivation. The general self-esteem construct has been the most studied (Table 2). There is both quantitative and qualitative evidence of some benefits of ECPAP. For the motivation construct, there is qualitative evidence of positive effects. For many of the constructs studied, no effect was observed. Jago et al. (2015) is the only authors whose ECPAP lead to smaller increases in psychosocial measures in the intervention group than in the control group.

### Risk of bias

The results of the risk bias assessment are presented in Appendix B. The overall quality of the studies was moderate, with an average score of 58%. Seven out of the 11 studies assessed had a score higher than 50%. The overall risk of reporting bias was rather low. Most of the methodological flaws were in the randomisation, blinding and power calculation.

### Recommendations for implementation and success

The recommendations came from different types of studies. Two were process evaluation studies (Robbins et al., 2016; Sebire et al., 2016), one formative (Rajan & Basch, 2012) evaluation study, one study assessed specifically implementation issues (Edwards et al., 2016) and eight intervention studies providing recommendations based on their experience (Betul, 2014; Gatz & Kelly, 2018; Jago et al., 2015; Jago et al., 2012; Kargarfard et al., 2012; Martin & Fairclough, 2008; Robbins et al., 2012; Watson et al., 2000). These findings have been regrouped into categories, as presented in Table 3. Among the recommendations, it should be highlighted that five studies proposed ways to enhance ECPAP designed for adolescent girls so that they improve MVPA and four studies included recommendations for increasing participation and attendance.

**Table 3.** Recommendations for implementation and success of extra-curricular physical activity programs, by program’s desired effect

|  |  |
| --- | --- |
| **Participation / attendance** |  |
| - House the program in the school (Watson et al., 2000)<br>- Mimic usual school provision (Edwards et al., 2016)<br>- Use non-schools personal to plan and conduct the program (Watson et al., 2000)<br>- Engage parents in providing support (Edwards et al., 2016)<br>- Use cooperative activities (interactions) (Watson et al., 2000)<br>- Use rewards that conduct to feelings of competence and autonomy (Robbins et al., 2016)<br>- Inform girls they have to attend for the total duration of the program (Robbins et al., 2016) | - Allow open enrollment (Edwards et al., 2016)<br>- Begin with activities requiring minimal skills (Watson et al., 2000)<br>- Offer only 1 session per week (Robbins et al., 2016)<br>- Maintain fun as the primary focus (Watson et al., 2000)<br>- Create a non-competitive environment (Watson et al., 2000)<br>- Consider:<br>o Transportation (Robbins et al., 2012)<br>o Other school events and activities (Edwards et al., 2016)<br>o Cost and resources needed (Robbins et al., 2016) |
| **Participants’ enjoyment** |  |
| - Ensure variety in session (e.g. different types of dance) (Edwards et al., 2016)<br>- Favor smaller group size (e.g. lower than 25 participants) (Sebire et al., 2016) | - Include group work (Edwards et al., 2016) |
| **Moderate-to-vigorous PA** |  |
| - Use aerobic warm-up (Martin & Fairclough, 2008)<br>- Reduce non-active time (R. Jago et al., 2012)<br>- Increase session intensity with participants’ improvement (R. Jago et al., 2012)<br>- Facilitate high-intensity sessions (e.g. dance sessions) (Russell Jago et al., 2015)<br>- Assess possible reduction in PA (e.g. active transportation) (Russell Jago et al., 2015) | - Involve program leader with knowledge on PA (Martin & Fairclough, 2008)<br>- Broaden activity type (e.g. different types of dance) (R. Jago et al., 2012)<br>- Involve both girls and their mother in PA session (Kargarfard et al., 2012)<br>- Create activity-friendly environment (e.g. encouragement from leaders and engagement of leaders in PA) (Robbins et al., 2016) |
| **Self-esteem** |  |
| - Use recreation activities; i.e. relaxing and entertaining activities (Betul, 2014) | - Ensure participation is voluntary (Betul, 2014) |
| **Skills progression** |  |
| - Broaden activity type (e.g. different types of dance) (R. Jago et al., 2012) |  |
| **Enrollment of the least active girls** |  |
| - Start recruitment in late September (Russell Jago et al., 2015) |  |
| **Overall success of the programs** |  |
| - Implement strategies to improve intervention fidelity, such as comprehensive professional development for teachers/coaches (Dewar et al.) | - Include environmental changes to support health behaviour changes (Dewar et al.) |
| - Recruit a bank of reserve instructors (Edwards et al., 2016) | - Set challenging goals for participants (Gatz & Kelly, 2018) |
| **Successful implementation of a multi-approach program** |  |
| - Teach simple lessons with only few games (Rajan & Basch, 2012) | - Include enough time to process the health topic discussed as a group (Rajan & Basch, 2012) |
| - Incorporate lessons that address only few sub-topics (Rajan & Basch, 2012) | - Teach lessons with clear and specific directives and learning objectives (Rajan & Basch, 2012) |
| - Teach lesson content that matches the participants' development stage (Rajan & Basch, 2012) | - Allow enough time for quiet and individual self-reflection (Rajan & Basch, 2012) |
| - Ensure the coaches are equipped with resources on each health topic (Rajan & Basch, 2012) | - Provide opportunities for the coaches to role-play or practice the lessons (Rajan & Basch, 2012) |
| - Provide coaches with information on the nature of emotional and physical developmental changes during adolescence (Rajan & Basch, 2012) | - Ensure the program takes place in a space free from distractions (Rajan & Basch, 2012) |
| - Plan for a large-enough indoor space as safe and dry alternative during inclement weather (Rajan & Basch, 2012) | - |
| PA: Physical activity |  |

## Discussion

This is the first systematic review aiming at evaluating the specific effects of ECPAP on both physical and psychological outcomes in high-school girls. The body of evidence reviewed presents a moderate risk of bias and showed very little improvement in PA levels and fitness markers. It also showed mixed results regarding the effect of ECPAP on body composition, with benefits observed only in shorter programs. Self-esteem was the most studied psychosocial construct, but evidence showed inconsistent results. Recommendations for successful implementation were mostly about improving program’s participation and MVPA.

This review gathered robust data on MVPA but showed limited improvements in response to ECPAP. Data were objectively measured and came from studies presenting a moderate risk of bias. Consequent to our findings, other reviews found mitigated improvements in MVPA levels with PA programs run in different contexts and settings (Camacho-Minano et al., 2011; Pearson et al., 2015; Voskuil et al., 2017). Hence, it appears that increasing MVPA using ECPAP seems challenging, especially if PA sessions are run only once a week. Considering that MVPA generally decreases during adolescence (Dumith et al., 2011), programs that help keep adolescent girls active could be considered as successful programs (Stone, McKenzie, Welk, & Booth, 1998). ECPAP included in this review lasted up to 12 months. Hence, one could think that keeping participants as active as they were at baseline is a positive outcome. The lack of change in MVPA could explain why most ECPAP did not lead to improved fitness markers. Exercise intensity might have been too small to affect them. Only one intervention led to increased fitness markers (Kargarfard et al., 2012). The differences between the interventions’ duration, frequency and attendance to PA sessions were not large enough to explain this observation. For instance, all ECPAP assessing fitness markers included between 2 to 5 sessions per week and compliance was higher than 60%, except for the Girls on the Move program.(Robbins et al., 2012) The explanation may reside in the difference in the populations studied. The CASPIAN study enrolled slightly older participants, had sufficient power and included the participation of the participants’ mothers as a motivational component. The effect of ECPAP on fitness markers should be verified with a larger number of studies for each fitness marker.

Perhaps current ECPAP do not help increase MVPA and fitness markers, but could improve health by influencing other lifestyle habits, such as time spent in sedentary activities and sleep habits. The 24h-movement guidelines gathers including recommendations on screen time and sleep durations for the youth, highlighted the importance for health of other movement behaviors than MVPA.(Tremblay et al., 2016) In the present review, only one research group assessed the effect of their ECPAP on a marker of sedentary time and observed a decrease.(Dewar et al., 2014; Lubans et al., 2012) None reported the sleep habits of their participants. There is some evidence that adolescents who participate in organised sports show lower risk of engaging in unhealthy behaviours, such as screen-based activities and short sleep duration.(Torstveit, Johansen, Haugland, & Stea, 2018) It could then be interesting to evaluate whether ECPAP can affect the health of participants by influencing sedentary activities and sleep habits.

From the body of evidence reviewed, it seems that shorter ECPAP (Kargarfard et al., 2012; Kelishadi et al., 2010; Yoshimoto et al., 2016) affected body composition more than longer duration ones.(Lubans et al., 2012; Robbins et al., 2012) Similarly, Voskuil et al. (2017) reviewed PA interventions for girls and all nine studies lead to no change in adiposity markers lasting more than 16 months. (Voskuil et al., 2017) Three out of five interventions that affected body mass index or percent body fat lasted 12 weeks, which is the same benchmark observed in this review. The length of the programs could offset the influence of the nutritional and motivational aspects of the multicomponent ECPAP on body composition. In fact, programs that included nutritional(Lubans et al., 2012) and motivational (Lubans et al., 2012; Robbins et al., 2012) aspects were also the longest. In this regard, most weight management programs show their greatest impact within the first few weeks, independent of the nature of the intervention. (Hall & Kahan, 2018)

This review is the first to evaluate the impact of PA programs on psychosocial outcomes among adolescent girls. This work shows some evidence that ECPAP could improve constructs related to self-esteem, physical competence and motivation (Table 2). The studies that showed such benefits had the lowest rating for methodological quality but this may simply come from the evaluation tool used to assess the risk of bias.(Downs & Black, 1998) Though this tool was built to evaluate both randomised and non-randomised studies, it might not be the best suited for qualitative studies. This tool has been chosen to assess best the majority of the studies and the majority of them used, at least in part, quantitative measures. Many constructs have been assessed in response to ECPAP, but most of them were evaluated as secondary objectives of the studies. Positive effects have been shown by studies assessing psychosocial outcomes as their primary objective. One could think that these studies were better suited to observe such outcomes. One study reported lower levels of motivation and satisfaction of participants’ need for competency and relatedness. (Jago et al., 2015) These observations were inconsistent with the authors’ hypotheses and they suggested that the timing of the last measure coinciding with a competition period could explain the observations, at least in part.

This section aims to provide concrete and useful tools to implement successful programs based on analyses that were discussed previously. These recommendations and implementations strategies (Table 3) were chosen according to their transferability and their extensive field of usage. They have been regrouped in the following bullet points.

▪ Participation/attendance and participants enjoyment: some authors highlighted the importance to allow open enrollment in the program and to host the program at school to ease participation. To maximise the enrollment of the least active girls, it is suggested to start recruiting them in late September, after the registration for organised sports. Engagement of parents by supporting their children and the use of activities that require minimal skill sets are also pointed out points in keeping participation high. Maintain fun as the central focus was a recurring theme. It was also important to ensure variety between sessions and to favour smaller groups per session (e.g. lower than 25 participants) to potentiate participants’ enjoyment.
▪ How to expand MVPA, self-esteem and skills progression: each session and the whole program should involve qualified personnel that have sufficient knowledge regarding PA. They should also be able to create a sustainable activity-friendly environment that can include encouragement from leaders and engagement of leaders in PA. To increase MVPA, periods of inactivity during sessions ought to be reduced and possible PA reduction factors (e.g. active transportation) should be assessed. Ensuring that participation is totally voluntary and the use of various activities are the key factors in increasing self-esteem and skills progression.
▪ Overall success of the program and successful implementation of a multi-approach program: to improve their success rate, programs should include environmental changes that elicit health behavioural changes. They should also set challenging goals for participants and provide a distraction-free environment. It is advised also to include simple lessons that address few sub-topics. Lessons should be taught with clear/specific directives on the learning objectives and should match the participants’ development stage.

### Study strengths and limitations

One strength of this review is the analysis of the methodological quality of studies describing the vast physiological and psychological effects of their ECPA (Appendix B). The overall risk of reporting bias was low. However, lack of power was observed in most studies. This could explain why many studies reported no significant differences. In addition, many of the included studies were non-randomised because of the nature of the interventions evaluated. In these situations, the subjects’ participation was based on their willingness to register for the ECPA; and therefore, induced some selection bias. The ECPA often aimed at increasing PA levels among girls who were the least active but authors often failed to recruit them.(Jago et al., 2015) The study of ECPAP created specifically for girls is still at an early stage. To improve the quality of the programs and research in the field, this review gathers the authors’ implementation recommendations, which is another strength of this study. Although some of the recommendations presented in Table 3 may not be suited for all types of ECPAP, researchers and program organisers can learn from experiences.

## Conclusion

This systematic review aimed to assess the effects of ECPAP on high-school girls. From the moderate quality of evidence systematically assessed, it seems that ECPAP does not lead to clear improvements in PA levels and fitness markers. Future studies should assess their effects on other health-related habits such as the reduction of sedentary time and sleep duration. There is some evidence that ECPAP could improve constructs related to self-esteem; however, more studies evaluating psychosocial outcomes as primary outcomes are recommended to obtain a clearer understanding. This review gathers recommendations to improve the efficacy of future ECPAP. The recurring shortcomings for which the authors provided advice were related to PA levels and MVPA as well as the increase in attendance and participation.

## Data Availability

All data referred to in this systematic review are available and published in peer-reviewed journals.

## Abbreviations

ECPAP: Extra-curricular physical activity programs
MVPA: Moderate-to-vigorous physical activity
PA: Physical activity

## Acknowledgments

The authors acknowledge the contribution of Denis Arvisais in the literature search. Marie-Eve Mathieu holds a *Fonds de Recherche en Santé du Québec* Junior 1 salary award.

## Appendix A PRISMA checklist

| Section/topic | # | Checklist item | Reported on page # |
| --- | --- | --- | --- |
| **TITLE** |  |  |  |
| Title | 1 | Identify the report as a systematic review, meta-analysis, or both. | Title page |
| **ABSTRACT** |  |  |  |
| Structured summary | 2 | Provide a structured summary including, as applicable: background; objectives; data sources; study eligibility criteria, participants, and interventions; study appraisal and synthesis methods; results; limitations; conclusions and implications of key findings; systematic review registration number. | Abstract |
| **INTRODUCTION** |  |  |  |
| Rationale | 3 | Describe the rationale for the review in the context of what is already known. | Introduction |
| Objectives | 4 | Provide an explicit statement of questions being addressed with reference to participants, interventions, comparisons, outcomes, and study design (PICOS). | Introduction |
| **METHODS** |  |  |  |
| Protocol and registration | 5 | Indicate if a review protocol exists, if and where it can be accessed (e.g., Web address), and, if available, provide registration information including registration number. | NA |
| Eligibility criteria | 6 | Specify study characteristics (e.g., PICOS, length of follow-up) and report characteristics (e.g., years considered, language, publication status) used as criteria for eligibility, giving rationale. | Methods |
| Information sources | 7 | Describe all information sources (e.g., databases with dates of coverage, contact with study authors to identify additional studies) in the search and date last searched. | Methods |
| Search | 8 | Present full electronic search strategy for at least one database, including any limits used, such that it could be repeated. | Methods |
| Study selection | 9 | State the process for selecting studies (i.e., screening, eligibility, included in systematic review, and, if applicable, included in the meta-analysis). | Methods |
| Data collection process | 10 | Describe method of data extraction from reports (e.g., piloted forms, independently, in duplicate) and any processes for obtaining and confirming data from investigators. | Methods |
| Data items | 11 | List and define all variables for which data were sought (e.g., PICOS, funding sources) and any assumptions and simplifications made. | Methods |
| Risk of bias in individual studies | 12 | Describe methods used for assessing risk of bias of individual studies (including specification of whether this was done at the study or outcome level), and how this information is to be used in any data synthesis. | Methods |
| Summary measures | 13 | State the principal summary measures (e.g., risk ratio, difference in means). | Methods |
| Synthesis of results | 14 | Describe the methods of handling data and combining results of studies, if done, including measures of consistency (e.g., $I^2$) for each meta-analysis. | Methods |
| Risk of bias across studies | 15 | Specify any assessment of risk of bias that may affect the cumulative evidence (e.g., publication bias, selective reporting within studies). | Methods |
| Additional analyses | 16 | Describe methods of additional analyses (e.g., sensitivity or subgroup analyses, meta-regression), if done, indicating which were pre-specified. | NA |
| **RESULTS** |  |  |  |
| Study selection | 17 | Give numbers of studies screened, assessed for eligibility, and included in the review, with reasons for exclusions at each stage, ideally with a flow diagram. | Results |
| Study characteristics | 18 | For each study, present characteristics for which data were extracted (e.g., study size, PICOS, follow-up period) and provide the citations. | Results |
| Risk of bias within studies | 19 | Present data on risk of bias of each study and, if available, any outcome level assessment (see item 12). | Results |
| Results of individual studies | 20 | For all outcomes considered (benefits or harms), present, for each study: (a) simple summary data for each intervention group (b) effect estimates and confidence intervals, ideally with a forest plot. | Results |
| Synthesis of results | 21 | Present results of each meta-analysis done, including confidence intervals and measures of consistency. | NA |
| Risk of bias across studies | 22 | Present results of any assessment of risk of bias across studies (see Item 15). | Results |
| Additional analysis | 23 | Give results of additional analyses, if done (e.g., sensitivity or subgroup analyses, meta-regression [see Item 16]). | NA |
| **DISCUSSION** |  |  |  |
| Summary of evidence | 24 | Summarize the main findings including the strength of evidence for each main outcome; consider their relevance to key groups (e.g., healthcare providers, users, and policy makers). | Discussion |
| Limitations | 25 | Discuss limitations at study and outcome level (e.g., risk of bias), and at review-level (e.g., incomplete retrieval of identified research, reporting bias). | Discussion |
| Conclusions | 26 | Provide a general interpretation of the results in the context of other evidence, and implications for future research. | Discussion |
| **FUNDING** |  |  |  |
| Funding | 27 | Describe sources of funding for the systematic review and other support (e.g., supply of data); role of funders for the systematic review. | Acknowledgments |

NA: not applicable

## Appendix B Methodological quality of studies presenting the effects of extra-curricular physical activity programs

| Criteria | Score by study |  |  |  |  |  |  |  |  |  |  |
| --- | --- | --- | --- | --- | --- | --- | --- | --- | --- | --- | --- |
|  | Waldron, 2007 | Martin & Fairclough, 2008 | Kelishadi et al., 2010 | Kargafard et al., 2012 | Lubans et al., 2012 | Robbins et al., 2012 | Betul, 2014 | Dewar et al., 2014 | Jago et al., 2015 | Yoshimoto et al., 2016 | Gatz & Kelly, 2018 |
| **Reporting** |  |  |  |  |  |  |  |  |  |  |  |
| 1. Is the hypothesis/aim/objective of the study clearly described? | 1 | 1 | 0 | 1 | 1 | 1 | 1 | 1 | 1 | 1 | 1 |
| 2. Are the main outcomes to be measured clearly described in the Introduction or Methods section? | 1 | 1 | 1 | 1 | 1 | 1 | 1 | 1 | 1 | 1 | 1 |
| 3. Are the characteristics of the patients included in the study clearly described? | 1 | 0 | 0 | 1 | 1 | 1 | 0 | 1 | 1 | 0 | 0 |
| 4. Are the interventions of interest clearly described? | 1 | 1 | 1 | 1 | 1 | 1 | 1 | 1 | 1 | 1 | 1 |
| 5. Are the distributions of principal confounders in each group of subjects to be compared clearly described? | 1 | 0 | 1 | 1 | 0 | 0 | 0 | 0 | 1 | 1 | 1 |
| 6. Are the main findings of the study clearly described? | 1 | 1 | 1 | 1 | 1 | 1 | 1 | 1 | 1 | 1 | 1 |
| 7. Does the study provide estimates of the random variability in the data for the main outcomes? | 1 | 1 | 1 | 1 | 1 | 1 | 1 | 1 | 1 | 1 | 1 |
| 8. Have all important adverse events that may be a consequence of the intervention been reported? | 1 | 0 | 0 | 0 | 1 | 0 | 0 | 1 | 1 | 1 | 0 |
| 9. Have the characteristics of patients lost to follow-up been described? | 1 | 1 | 1 | 1 | 1 | 1 | 1 | 1 | 1 | 1 | 0 |
| 10. Have actual probability values been reported (e.g. 0.035 rather than <0.05) for the main outcomes except where the probability value is less than 0.001? | 1 | 0 | 1 | 1 | 0 | 1 | 1 | 1 | 1 | 1 | 1 |
| **External validation** |  |  |  |  |  |  |  |  |  |  |  |
| 11. Were the subjects asked to participate in the study representative of the entire population from which they were recruited? | 0 | 0 | 1 | 1 | 0 | 1 | 0 | 0 | 1 | 0 | 0 |
| 12. Were those subjects who were prepared to participate representative of the entire population from which they were recruited? | 0 | 0 | 0 | 0 | 0 | 0 | 0 | 0 | 0 | 0 | 0 |
| 13. Were the staff, places, and facilities where the patients were treated, representative of the treatment the majority of patients receive? | 1 | 1 | 1 | 1 | 1 | 1 | 0 | 1 | 1 | 0 | 1 |
| Internal validation - bias |  |  |  |  |  |  |  |  |  |  |  |
| 14. Was an attempt made to blind study subjects to the intervention they have received? | 0 | 0 | 0 | 0 | 0 | 0 | 0 | 0 | 0 | 0 | 0 |
| 15. Was an attempt made to blind those measuring the main outcomes of the intervention? | 0 | 0 | 0 | 0 | 1 | 0 | 0 | 1 | 1 | 1 | 0 |
| 16. If any of the results of the study were based on “data dredging”, was this made clear? | 1 | 1 | 1 | 1 | 1 | 1 | 1 | 0 | 1 | 1 | 1 |
| 17. In trials and cohort studies, do the analyses adjust for different lengths of follow-up of patients, or in case-control studies, is the time period between the intervention and outcome the same for cases and controls? | 0 | 1 | 0 | 0 | 1 | 1 | 0 | 1 | 1 | 1 | 1 |
| 18. Were the statistical tests used to assess the main outcomes appropriate? | 1 | 1 | 1 | 1 | 1 | 1 | 1 | 1 | 1 | 1 | 1 |
| 19. Was compliance with the intervention/s reliable? | 0 | 1 | 1 | 1 | 1 | 0 | 0 | 1 | 0 | 1 | 0 |
| 20. Were the main outcome measures used accurate (valid and reliable)? | 1 | 1 | 1 | 1 | 1 | 1 | 1 | 1 | 1 | 1 | 1 |
| Internal validation – confounding (selection bias) |  |  |  |  |  |  |  |  |  |  |  |
| 21. Were the patients in different intervention groups (trials and cohort studies) or were the cases and controls (case-control studies) recruited from the same population? | 0 | 0 | 0 | 0 | 1 | 1 | 0 | 1 | 1 | 1 | 0 |
| 22. Were study subjects in different intervention groups (trials and cohort studies) or were the cases and controls (case-control studies) recruited over the same period of time? | 0 | 0 | 0 | 0 | 1 | 1 | 0 | 1 | 1 | 0 | 0 |
| 23. Were study subjects randomized to intervention groups? | 0 | 0 | 0 | 0 | 0 | 1 | 0 | 0 | 1 | 0 | 0 |
| 24. Was the randomized intervention assignment concealed from both patients and health care staff until recruitment was complete and irrevocable? | 0 | 0 | 0 | 0 | 0 | 0 | 0 | 0 | 1 | 0 | 0 |
| 25. Was there adequate adjustment for confounding in the analyses from which the main findings were drawn? | 0 | 0 | 0 | 0 | 0 | 0 | 0 | 1 | 1 | 0 | 0 |
| 26. Were losses of patients to follow-up taken into account? | 1 | 1 | 1 | 1 | 1 | 1 | 1 | 1 | 1 | 1 | 0 |
| Power |  |  |  |  |  |  |  |  |  |  |  |
| 27. * Did the study have sufficient power to detect a clinically important effect where the probability value for a difference being due to chance is less than 5%? Sample sizes have been calculated to detect a difference of x% and y%. | 0 | 0 | 0 | 0 | 0 | 1 | 0 | 0 | 1 | 0 | 1 |
| Total score ( /28) | 15 | 13 | 14 | 16 | 18 | 19 | 11 | 19 | 24 | 17 | 12 |
| Percentage (%) | 54 | 46 | 50 | 57 | 64 | 68 | 39 | 68 | 86 | 61 | 43 |

